# Screening Practice of Major Congenital Malformations and Associated Factors among Healthcare Professionals at Three Teaching Hospitals in Ethiopia: a cross-sectional study

**DOI:** 10.64898/2025.12.25.25343015

**Authors:** Eden Belay Tilahun, Fisseha Temesgen, Abraham Sisay Abie, Nardos Mulu Admasu, Yirgalem Teklebirhan, Nahom Desalegn Mekonnen, Tsedey Zekarias Mena, Tigist Workneh Leulseged

**Author notes:** corresponding author Eden Belay Tilahun.

## Abstract

**Background:** Congenital anomalies are among the leading causes of neonatal morbidity and mortality, particularly in low- and middle-income countries. Early detection through newborn screening improves outcomes, yet little is known about the screening practices of healthcare professionals (HCPs) in Ethiopia. In our setting, many children with congenital malformations are not identified at birth and present later, after being discharged from the birth facility.

**Methods:** A cross-sectional study was conducted from November 2022 to September 2023 among 163 health care professionals (HCPs) working in labor and delivery room and neonatal care units at three tertiary hospitals in Addis Ababa. Data on knowledge, attitude, and practice (KAP) regarding the screening of major congenital malformations were collected through structured questionnaires and direct observation. Descriptive statistics and binary logistic regression were performed using SPSS version 26, with statistical significance set at p < 0.05.

**Results:** Majority of HCPs have moderate to adequate knowledge (74.2%), and favorable attitude (69.3%). However, only 11% demonstrated a good screening practice to identify major congenital anomalies. Adequate knowledge was strongly associated with good screening practice (AOR = 30.8; 95% CI: 0.15–0.95; p < 0.001). Infants born after complicated pregnancies and male newborns were more likely to be screened.

**Conclusion:** Screening practice among HCPs remains significantly below the world health organization (WHO) recommendations potentially leading to preventable morbidity, mortality and disability. Strengthening provider training and implementing standardized screening protocols are essential to improve early detection and reduce complications associated with congenital anomalies.

## 1. Introduction

### 1.1 Background

Congenital anomalies, also known as birth defects, are structural, functional, or metabolic abnormalities that arise during intrauterine development. These anomalies may present at birth or later in life and contribute significantly to infant morbidity and mortality worldwide. Although the etiology of approximately half of congenital anomalies remains unknown, known risk factors include genetic mutations, chromosomal abnormalities, intrauterine infections, teratogenic exposures (e.g., radiation, certain drugs, and pollutants), and maternal conditions such as diabetes or nutritional deficiencies) (1–6).

Congenital anomalies are classified into **major** and **minor** categories. Major anomalies such as neural tube defects, gastrointestinal atresia, and orofacial cleft have serious medical, surgical, or cosmetic consequences and often require urgent intervention. Minor anomalies, though less severe, can be indicative of underlying syndromes or chromosomal abnormalities (2,7,8).

Newborn screening is a crucial public health strategy aimed at the early detection of congenital disorders. Studies have suggested that the initial newborn examination must be conducted by a qualified professional within 24hours of the life since the majority of anatomic malformations in resource limited settings are detectable at birth and may necessitate urgent intervention to ameliorate adverse outcomes. A second examination is performed at 6-8 weeks of age to identify abnormalities that develop or become apparent later. Effective screening requires thorough history-taking, physical examination, and where indicated, further diagnostic testing (2,4,9,10,11,12)

### 1.2 Statement of the Problem

Globally, congenital anomalies affect approximately 3% of live births and account for 20–30% of infant mortality. According to the World Health Organization (WHO), over 240,000 newborns die within the first 28 days of life annually due to birth defects. Additionally, congenital anomalies are responsible for an estimated 170,000 deaths among children aged 1 month to 5 years.

In low- and middle-income countries (LMICs), the health burden from congenital anomalies is disproportionately high due to limited access to preventive, diagnostic, and treatment services. Factors such as underreporting, lack of screening protocols, limited public awareness, and inadequate training of healthcare providers contribute to delayed diagnosis and suboptimal outcomes. In Ethiopia, the true prevalence of congenital anomalies remains unclear, but available hospital-based studies indicate a high incidence, compounded by delayed presentation and diagnosis.

### 1.3 Significance of the Study

Although most major congenital anomalies can be detected through careful clinical examination at birth, many infants in Ethiopia present late leading to delayed treatment and poor prognosis. This delay is further exacerbated by a lack of standardized screening tools, inadequate documentation, and insufficient training among healthcare providers.

This study is the first of its kind in Ethiopia to assess the knowledge, attitudes, and practices (KAP) of healthcare professionals involved in newborn care regarding screening for congenital malformations. By identifying gaps in practice and associated factors, the findings can inform policy changes, staff training initiatives, and development of national screening protocols. Ultimately, this research aims to improve early detection and timely management of congenital anomalies, reducing the burden of preventable neonatal morbidity and mortality.

## 2. Literature Review

In high-income countries, routine screening for congenital anomalies is an established component of neonatal care. However, in many LMICs, such screening is underdeveloped or non-existent. The lack of standardized protocols, insufficient healthcare provider training, and limited access to diagnostic tools contribute to delayed identification and increased morbidity and mortality in affected infant Parental responsibility for identifying newborns with birth abnormalities is disproportionately increased in the absence of precise recommendations for newborn screening (14).

A study in Pakistan revealed that over half of infants with anorectal malformations (ARMs) were diagnosed by non-medical personnel, despite being delivered in healthcare facilities. The median time to diagnosis was 48 hours, underscoring gaps in early neonatal assessment (8). Similarly, a study from Malawi reported that only 17% of neonates with ARMs were identified within the first week of life (7).

Another study, prospective observational one done India on Anorectal malformations comparing the timing of diagnosis with the outcome has found that nearly half (48%) of all neonates presenting with ARM had a delayed diagnosis (> 48 h after birth(10) which was found to be higher compared to reports from developed countries like by Wilson et al. (32%), and Turowski et al. (21.2%) (11,12)

In contrast, data from high-income settings demonstrate more timely detection. For example, in the UK, one study found that midwives conducted newborn examinations with comparable accuracy to physicians and achieved higher maternal satisfaction, highlighting the value of empowering non-physician providers in neonatal care (15).

Delayed diagnosis of congenital anomalies such as esophageal atresia and tracheoesophageal fistula (EA/TEF) is a significant contributor to poor outcomes in LMICs. Inadequate awareness, inappropriate use of contrast studies, and lack of early referral protocols exacerbate risks such as aspiration pneumonia. Evidence suggests that early identification and referral are essential to improving surgical outcomes for EA/TEF and similar conditions (13).

In Ethiopia, Mekonnen et al. (2020) reported only 0.35% of births were documented with congenital anomalies in 37 public facilities in Addis Ababa, a figure likely reflecting under diagnosis rather than true prevalence. Contributing factors included poor diagnostic capacity, lack of trained personnel, and absence of congenital anomaly surveillance systems (5).

Several studies from LMICs have shown that health professionals often lack adequate knowledge or confidence to screen for congenital anomalies. In Saudi Arabia, nearly half of surveyed family physicians reported not screening for hearing loss in children over the past five years (17). In Egypt, over half of neurosurgical nurses demonstrated poor competency in managing infants with central nervous system anomalies (18).

Recommendations from international bodies, including WHO and CDC, emphasize the importance of neonatal screening during the first 24 hours of life and again at discharge or within 10 days. Despite these recommendations, many facilities in LMICs do not adhere to such timelines due to logistical, institutional, and training-related barriers (16).

The literature highlights a clear need to strengthen screening protocols, expand training opportunities, and institutionalize routine neonatal examinations in LMICs. Without these improvements, delays in diagnosis and care will continue to result in preventable disability and death among infants with congenital anomalies

## 3. Objectives

### 3.1 General Objective

- To evaluate the practice of healthcare professionals in screening for major congenital anomalies among newborns in three tertiary hospitals in Addis Ababa, Ethiopia.

### 3.2 Specific Objectives

- To assess the level of practice among healthcare professionals regarding screening of congenital malformations in newborns.
- To identify factors associated with healthcare professionals’ screening practices for congenital anomalies.
- To determine the knowledge and attitude of healthcare professionals about screening of major congenital abnormalities

## 4. Methods

### 4.1 Study Area and Period

This multicenter study was conducted from November 2022 to September 2023 in Addis Ababa, Ethiopia. The city is home to sixteen public hospitals, and three were purposively selected for this study based on their provision of neonatal intensive care and pediatric surgical services: Tikur Anbessa Specialized Hospital (TASH), St. Paul’s Hospital Millennium Medical College (SPHMMC), and Menelik II Comprehensive Specialized Hospital.

### 4.2 Study Design

An institution-based, cross-sectional analytical study design was employed.

### 4.3 Population

#### 4.3.1 Source Population

All HCPs involved in neonatal care at the selected hospitals’ labor and delivery wards and neonatal intensive care units (NICUs).

#### 4.3.2 Study Population

All eligible and randomly selected HCPs working in the labor wards and NICUs of the three hospitals during the study period.

### 4.4 Inclusion and Exclusion Criteria

**Inclusion Criteria:** HCPs actively involved in neonatal care during the data collection period.

**Exclusion Criteria**: There were no exclusion criteria for this study; all eligible participants who met the inclusion criteria were enrolled.

### 4.5 Sample Size Determination

The sample size was calculated using a single population proportion formula, assuming a 50% prevalence of adequate screening practice (due to lack of prior data), 95% confidence interval, and 5% margin of error:

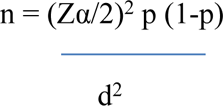

With the above the calculated sample size will be 384. Since the total population (health care professionals working in the delivery room and neonatal intensive care unit of the selected Hospitals is 242 n, which is less than 10,000, the adjustment formula is used.

The sample size after adjustment will be 148.46≈148. Adding a 10% non-response rate, the final sample size was adjusted to **163**.

### 4.6 Sampling Technique

A simple random sampling method was used. Proportional allocation was applied to ensure representation from each hospital (Figure 1)

- TASH: 54 participants
- SPHMMC: 77 participants
- Menilik II Hospital: 32 participants

**Figure 1:**
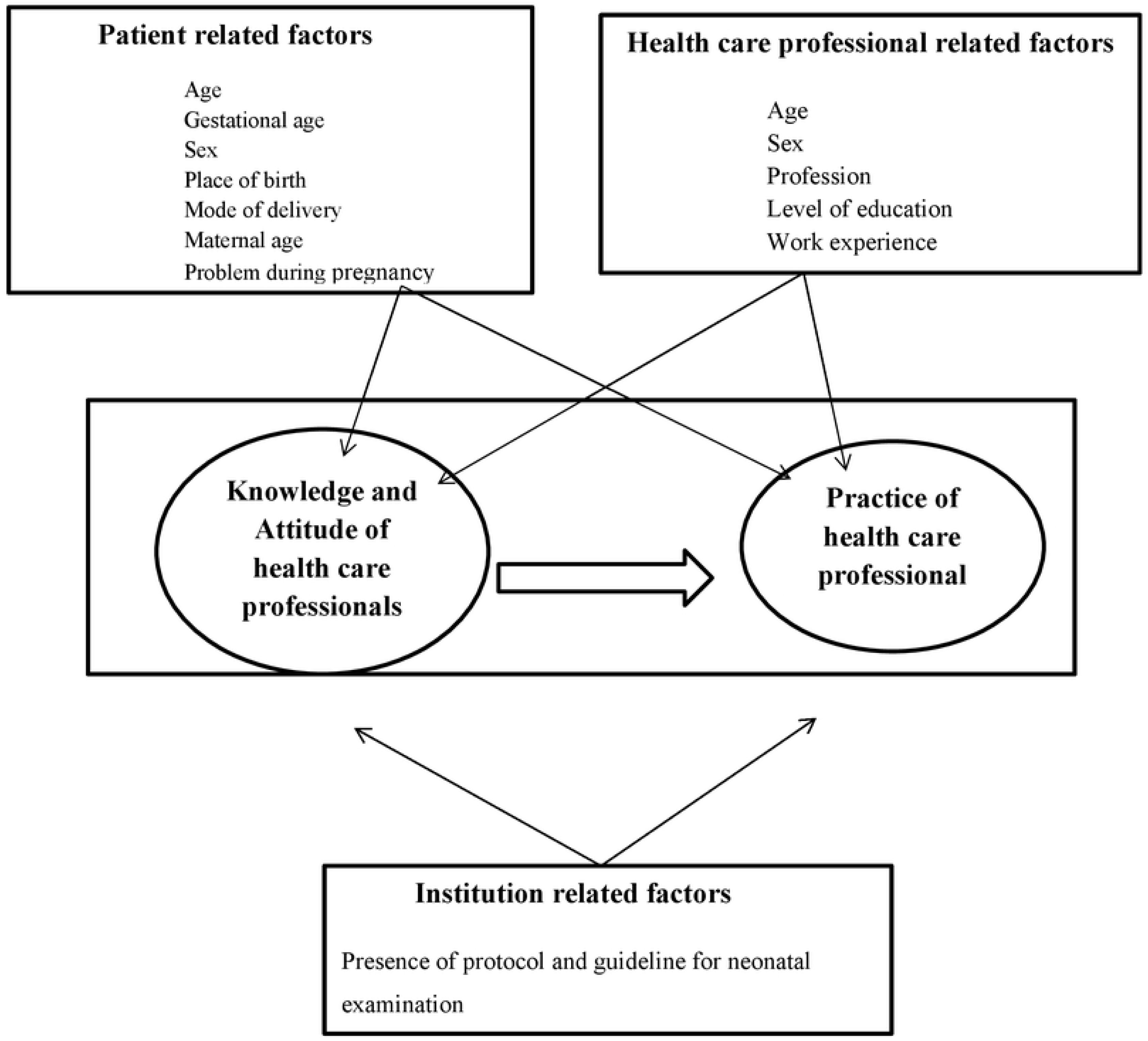
conceptual frame work.

**Figure 2:**
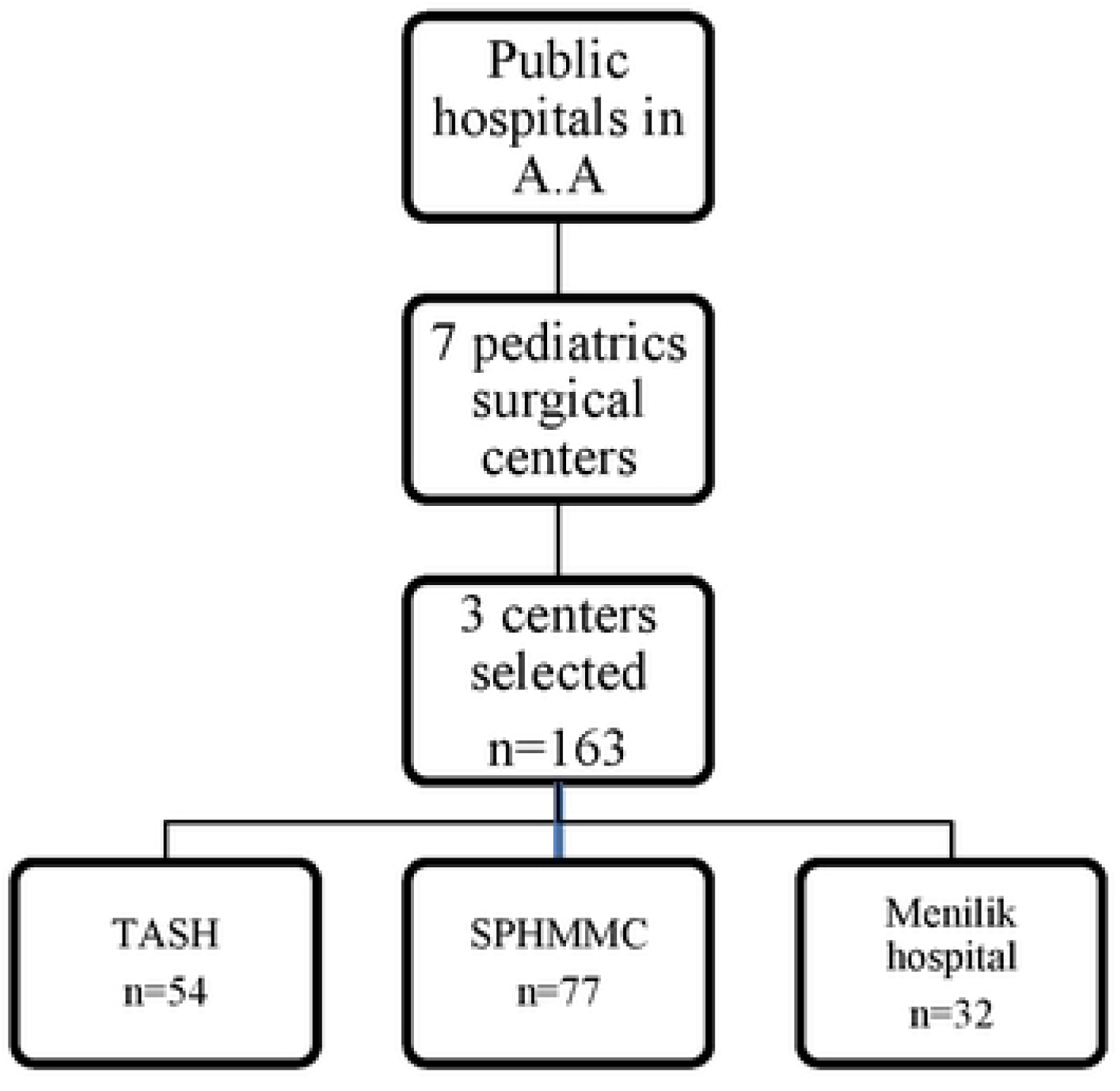
Sampling and procedure.

**Figure 3:**
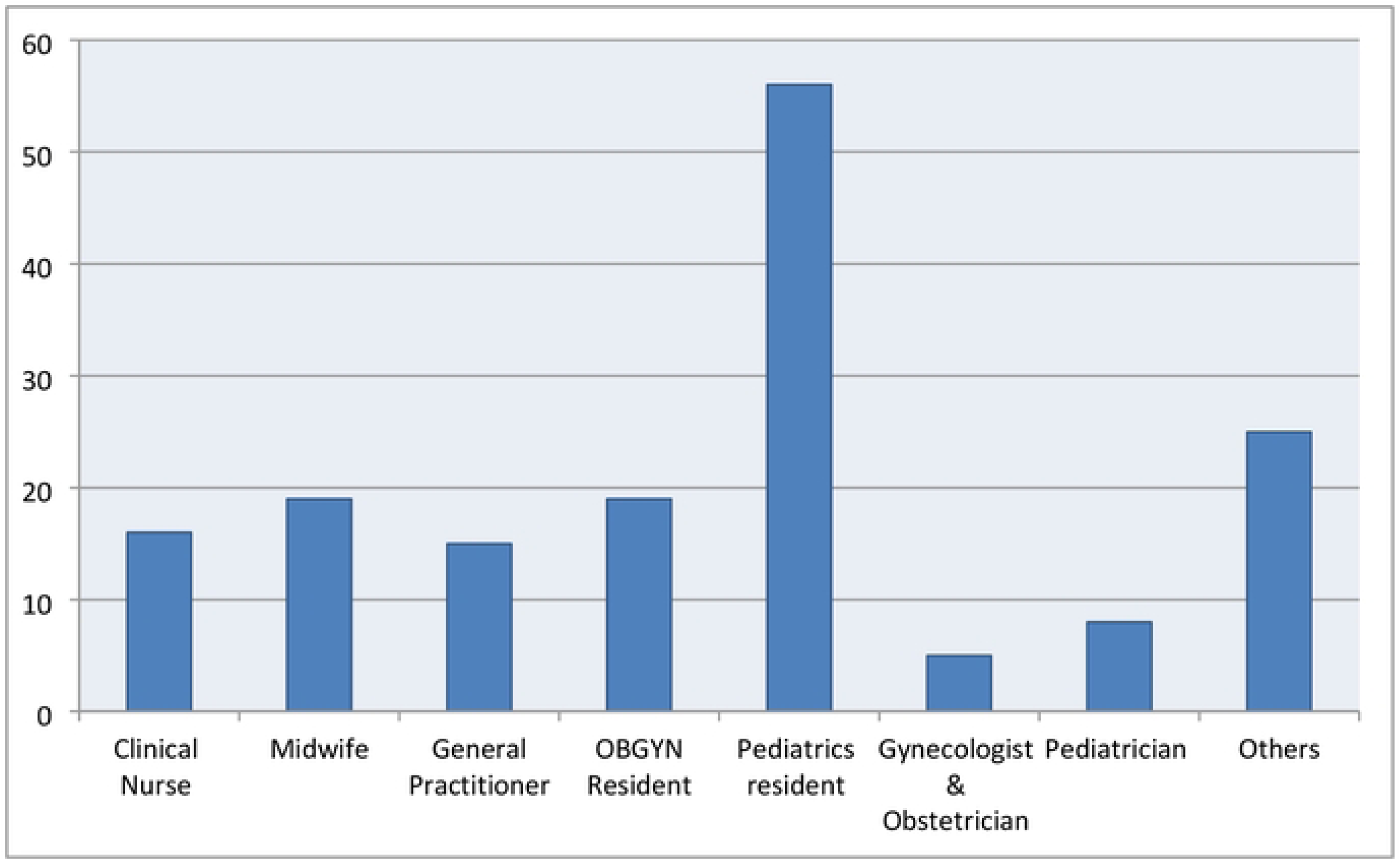
profession of participants.

### 4.7 Data Collection Tools and Procedures

Data was collected using a structured, pretested self-administered questionnaire and direct observation checklist. The questionnaire was adapted from WHO and CDC guidelines and covered demographics, knowledge, attitudes, and practices regarding congenital anomaly screening. Observations of HCP’s practices were conducted by trained data collectors.

### 4.8 Study Variables

- **Outcome Variable:** Practice of screening for congenital anomalies
- Independent Variables:

- **Patient-related factors:** sex, gestational age, place of birth, mode of delivery, maternal age, complications during pregnancy
- **Provider-related factors:** age, sex, profession, education level, work experience, prior training
- Knowledge and attitude scores
- **Institutional factors:** availability of screening protocols and tools Conceptual framework (Figure 1)

### 4.9 Operational Definitions

- **Good Practice:** ≥80% score on standardized practice checklist (based on WHO/CDC recommendations)
- **Adequate Knowledge:** ≥80% correct responses
- **Favorable Attitude:** ≥80% score on attitude questions
- **Congenital Malformations:** Major structural abnormalities with medical, surgical, or functional significance (e.g., neural tube defects, cleft lip/palate, anorectal malformations, undescended testis etc.)

### 4.10 Data Management and Analysis

Data was cleaned, coded, and entered into SPSS version 26. Descriptive statistics (frequencies, means, and medians) were computed. Binary logistic regression identified factors associated with good screening practice. Variables with p < 0.25 in bivariable analysis were included in multivariable regression. Statistical significance was set at p < 0.05.

### 4.11 Ethical Considerations

Ethical approval was obtained from the Institutional Review Board of Addis Ababa University College of health sciences school of medicine, department of surgery research ethics committee. Written informed consent was obtained from each participant. Confidentiality was maintained throughout the study.

## 5. Results

### 5.1 Sociodemographic Characteristics of Participants

A total of 163 healthcare professionals (HCPs) participated in the study, yielding a 100% response rate. Of the participants, 55.2% (n = 90) were male, and the median age was 28 years (IQR: 26–30). Most participants (71.2%) were under the age of 30.

Regarding professional roles, 46% were residents, followed by nurses and midwives (21.5%), general practitioners (9.2%) pediatricians (4.9%), obstetricians (3.1%), and others (16%) (Figure and Figure 4)

**Figure 4:**
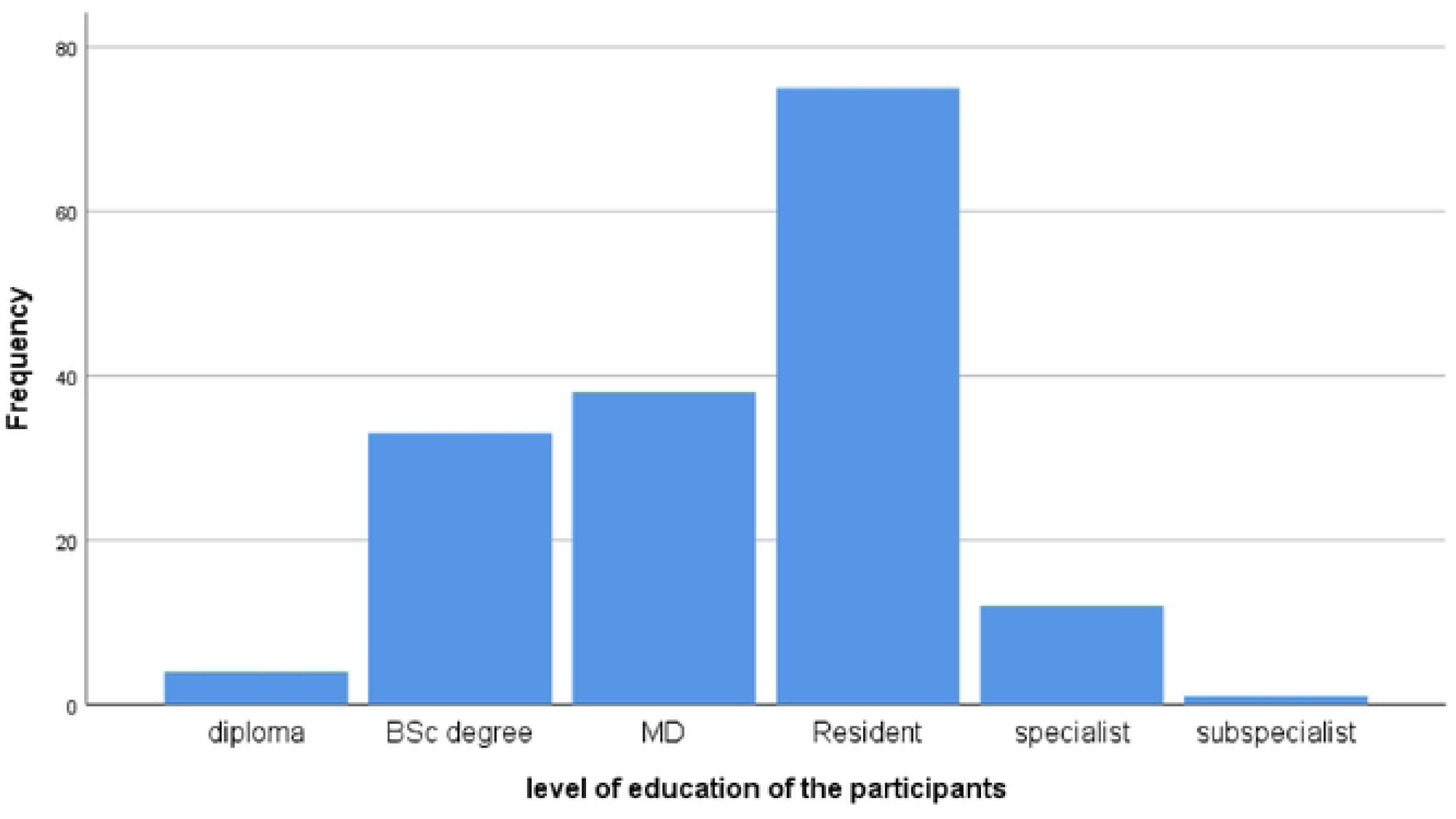
Level of education of the participants.

Approximately 77% of the participants had less than five years of work experience. Only 5.5% (n = 9) had received on-the-job training specifically focused on congenital malformation screening.

**Table 1:** Socio-demographic characteristics of HCPs.

| Characteristic |  | Number (%) |
| --- | --- | --- |
| Age group (years) | <30 | 115(70.6) |
|  | ≥30 | 48(29.4) |
| Sex | Male | 90(55.2) |
|  | Female | 73(44.8) |
| Profession | Clinical Nurse | 16(9.8) |
|  | Midwife | 19(11.7) |
|  | General Practitioner | 15(9.2) |
|  | OBGYN resident | 20(12.3) |
|  | Pediatrics resident | 54(32.5) |
|  | Obstetrician and gynecologist | 5(3.1) |
|  | Pediatrician | 8(4.9) |
|  | Others* | 26(16) |
| Level of education | Diploma | 7(4.3) |
|  | BSc Degree | 30(18.4) |
|  | MD | 38(23.3) |
|  | Resident | 75(46) |
|  | Specialist | 12(7.4) |
|  | Sub-Specialist | 1(0.6) |
| Work experience(years) | <5 | 125(76.7) |
|  | ≥5 | 38(23.3) |
| On-job Training on evaluation of congenital anomalies | No | 154(94.5) |
|  | Yes | 9(5.5) |
| Others*-Interns |  |  |

### 5.2 Knowledge, Attitude, and Practice (KAP) of Respondents

#### 5.2.1 Knowledge

The average knowledge score was 6.52 ± 1.48 out of a possible 10. Overall, 27.6% (n = 45) of participants demonstrated **adequate knowledge** (≥80%), while 46.6% had moderate knowledge and 25.8% had inadequate knowledge (Figure 5)

**Figure 5:**
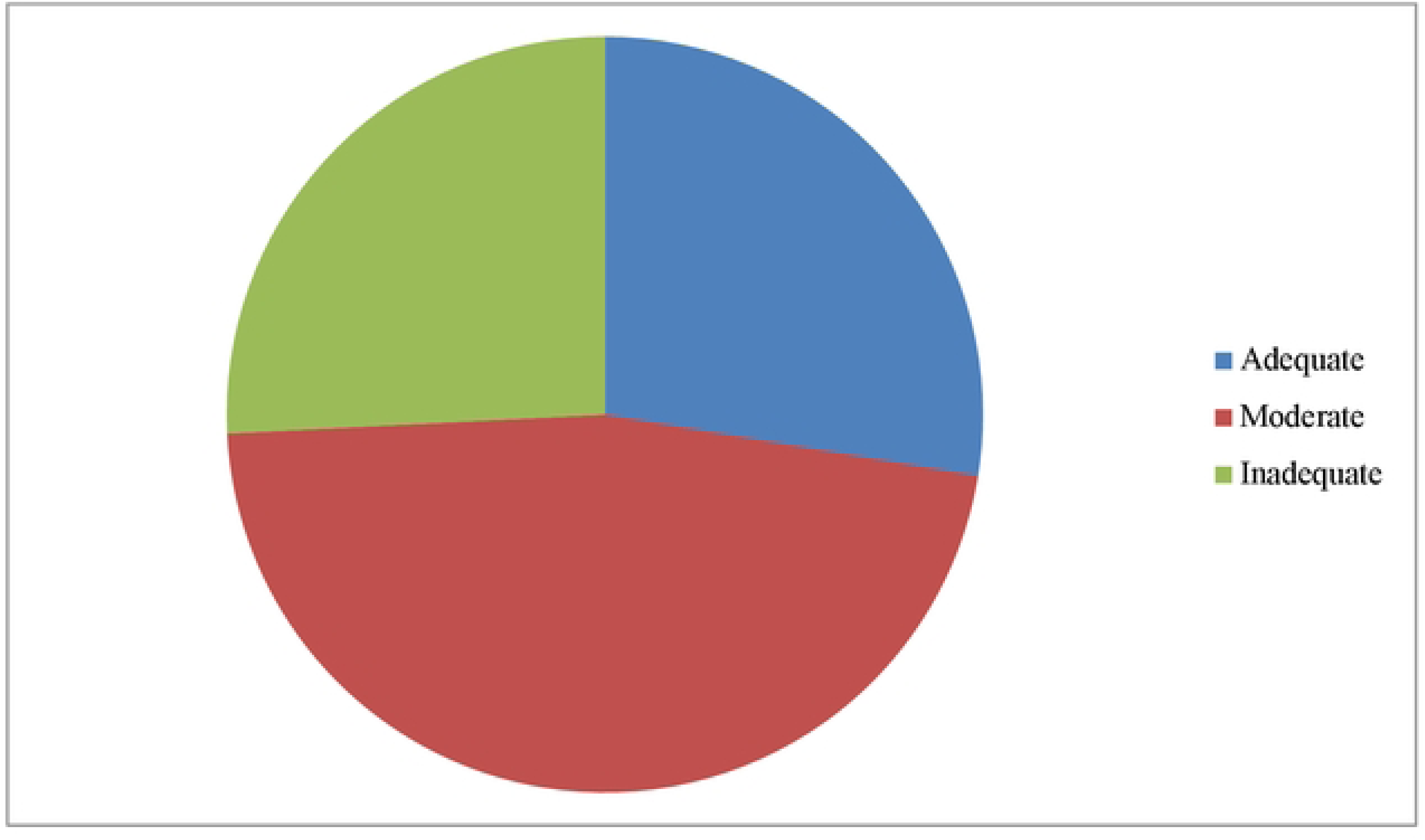
Knowledge level of the participants.

Knowledge gaps were noted regarding the timing of screening and management of specific anomalies. For example, only 58.9% correctly identified the ideal timing for newborn screening, and 33.7% correctly answered questions on managing abdominal wall defects.

**Table 2:**
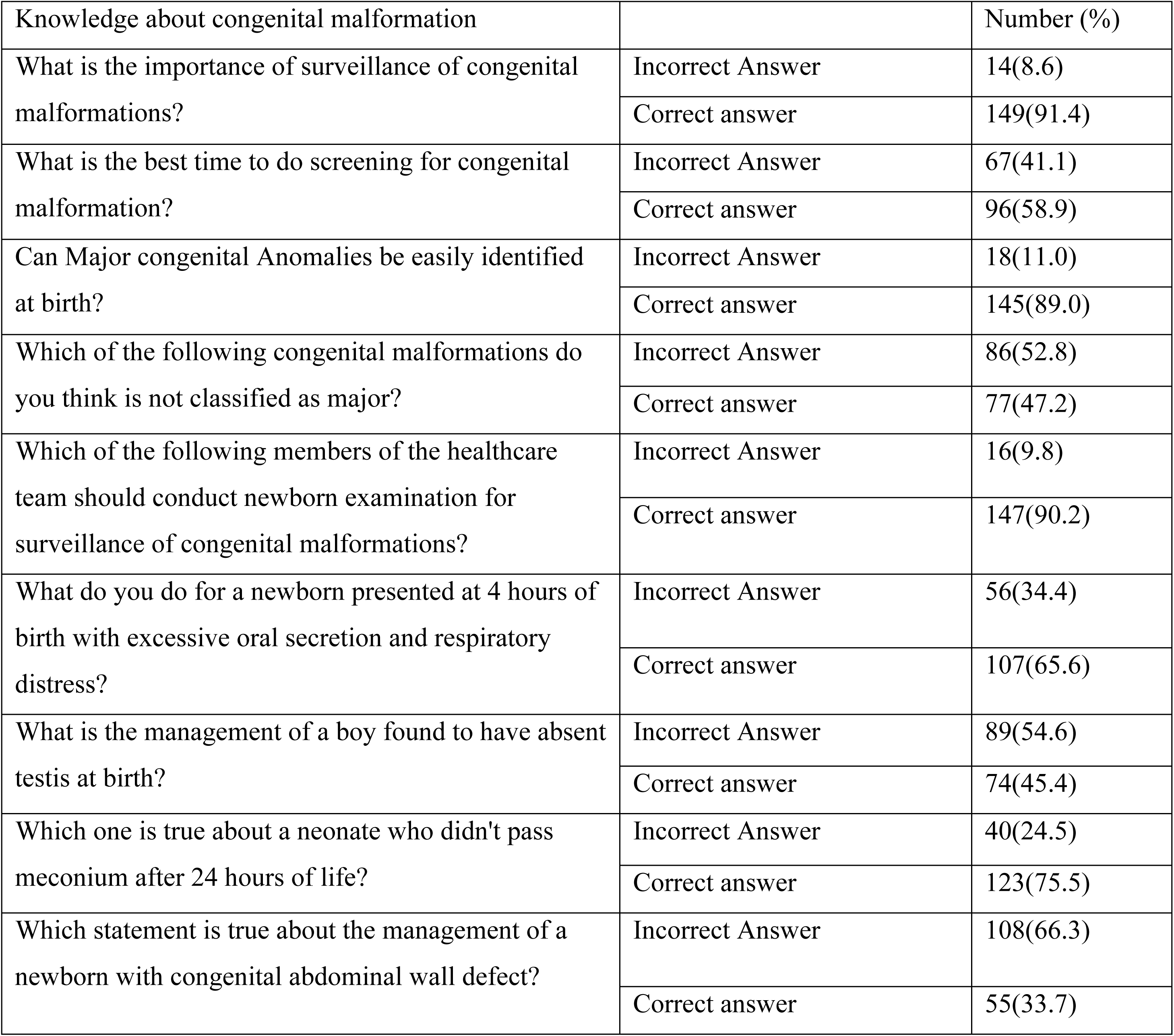
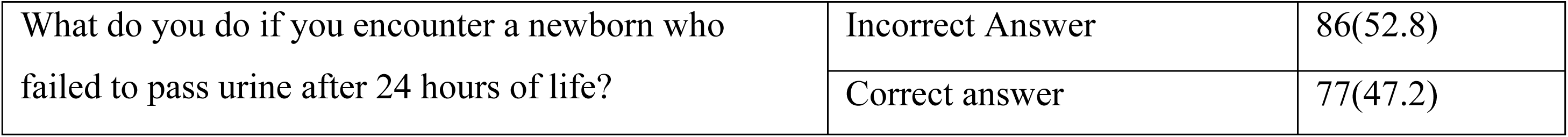
Knowledge of HCPs about congenital malformations.

#### 5.2.2 Attitude

The mean attitude score was 3.62 ± 0.30. A favorable attitude (≥80% score) was reported in 69.3% (n = 113) of participants. Notably, most respondents (77.3%) agreed that the lack of institutional guidelines hinders optimal screening. Similarly, 85.3% believed that inadequate staff training contributed to poor screening practices, while 68.1% cited workload and lack of time as major barriers (Figure 6).

**Figure 6:**
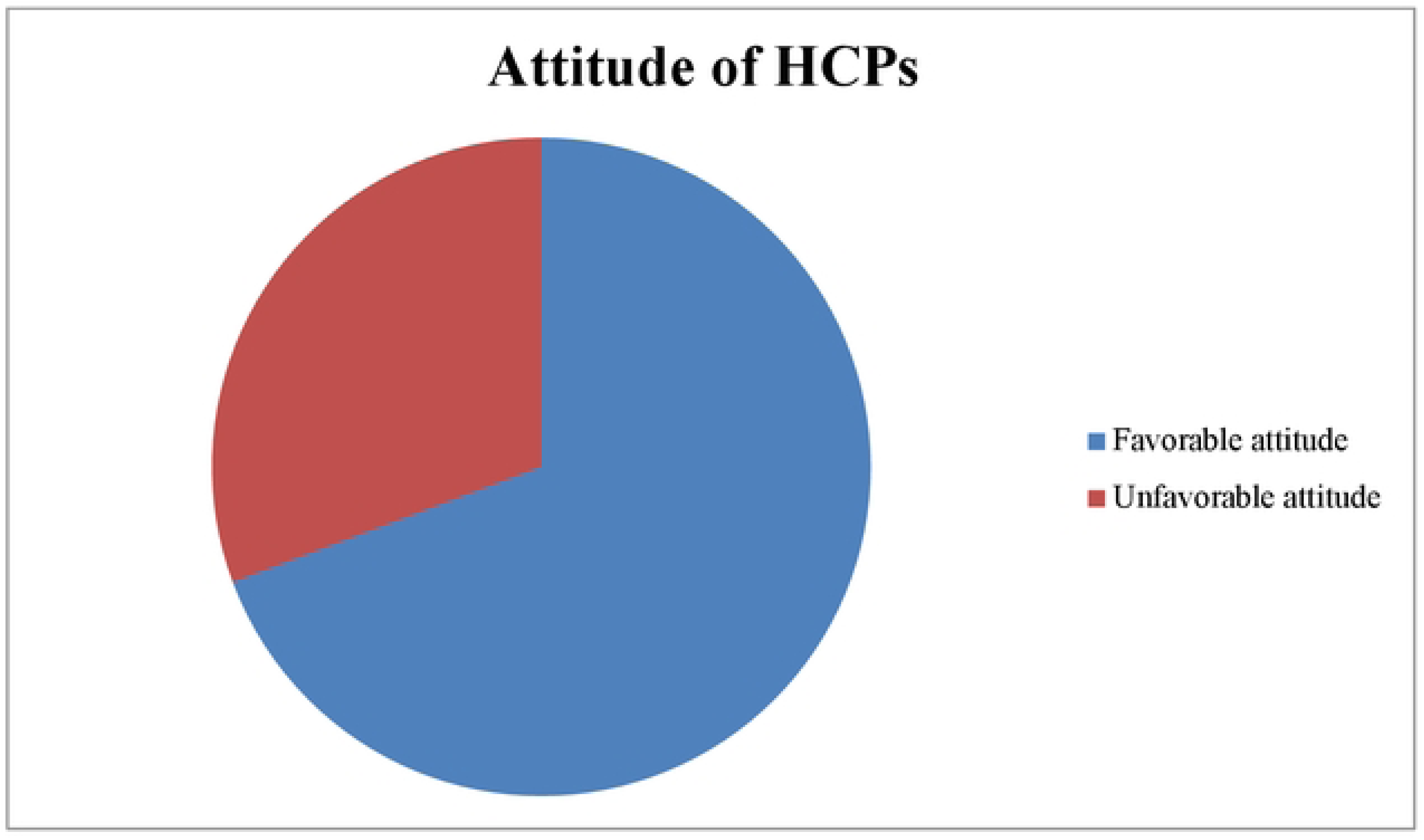
Attitude of HCPs regarding screening for congenital malformations.

**Table 3:**
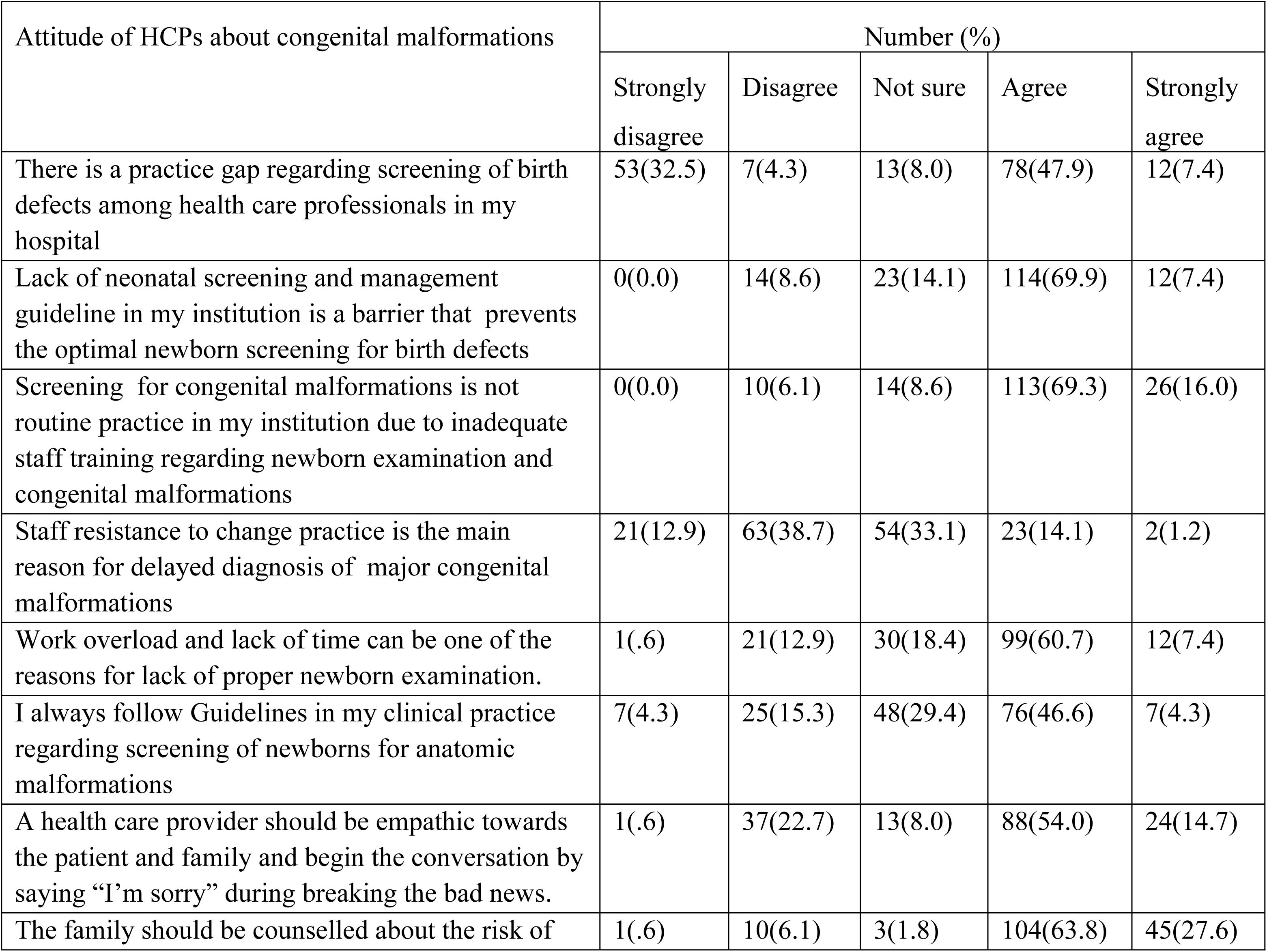
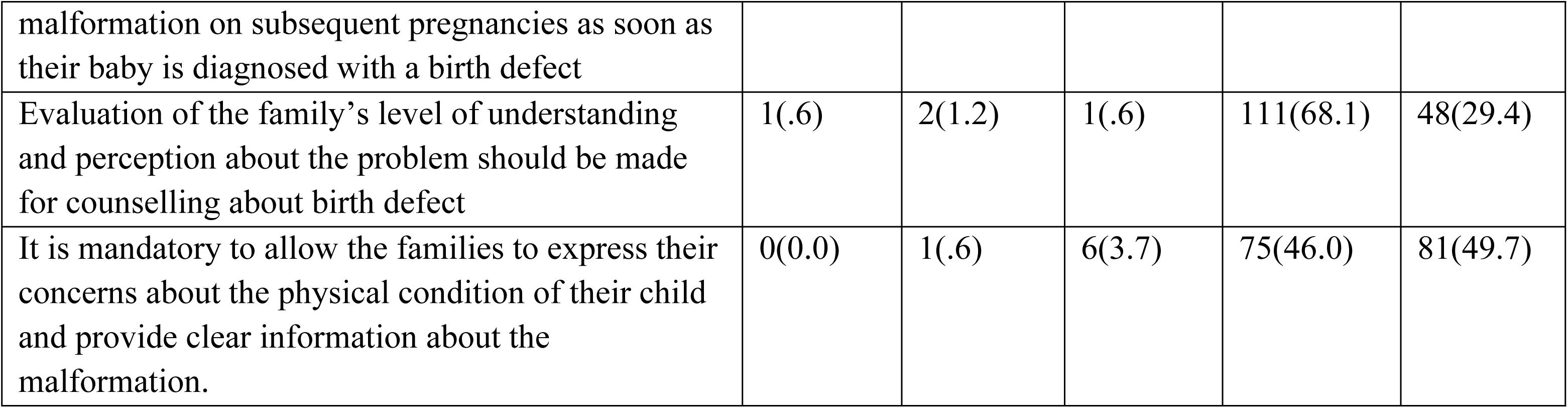
Attitude of HCPs about congenital malformations.

#### 5.2.3 Practice

The average practice score was 4.63 ± 1.79. Only 11% (n = 18) of HCPs demonstrated **good practice** (≥80% score), while the vast majority (89%) had poor screening practice (Figure 7).

**Figure 7:**
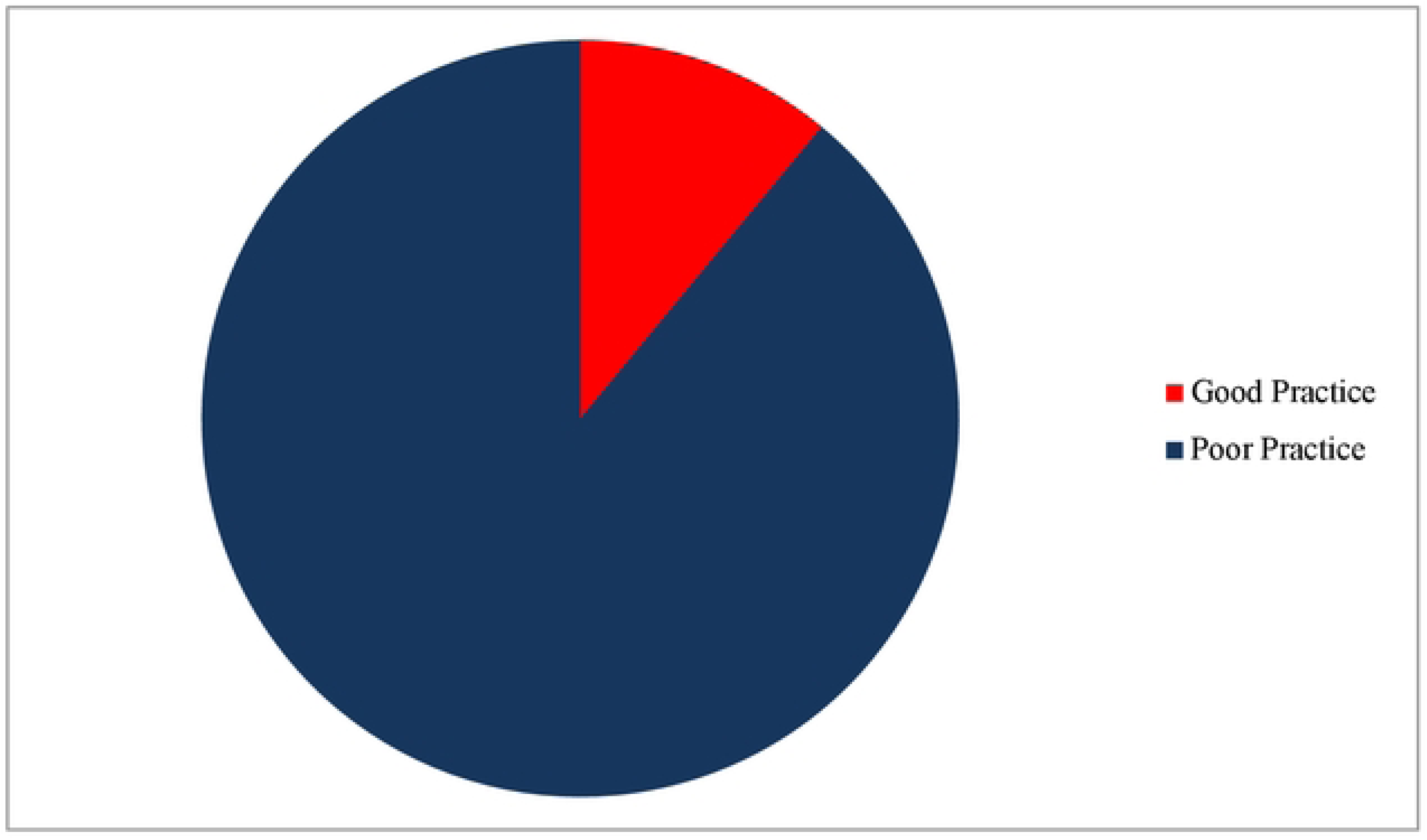
Practice of HCPs on screening major congenital malformations.

**Table 4:**
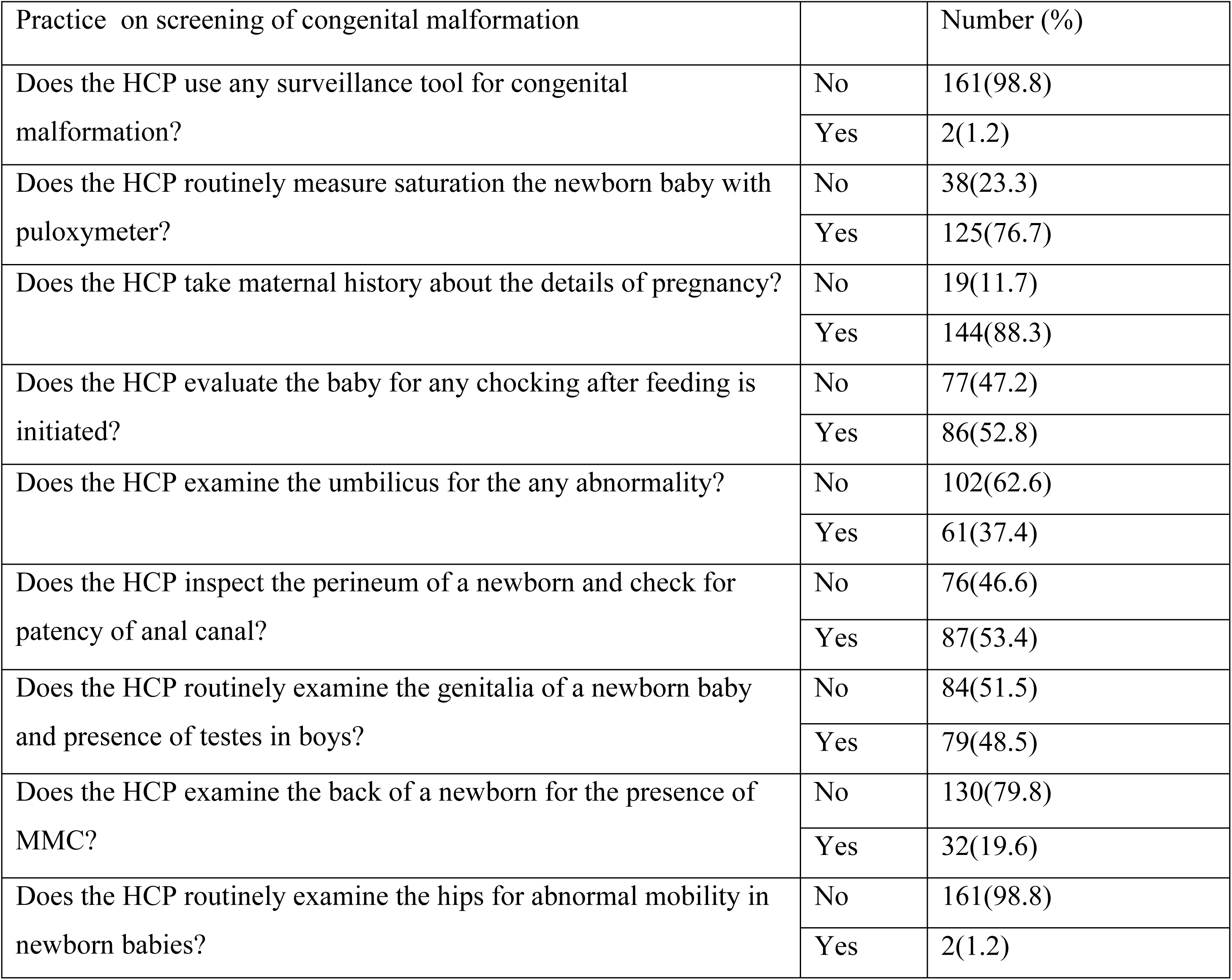
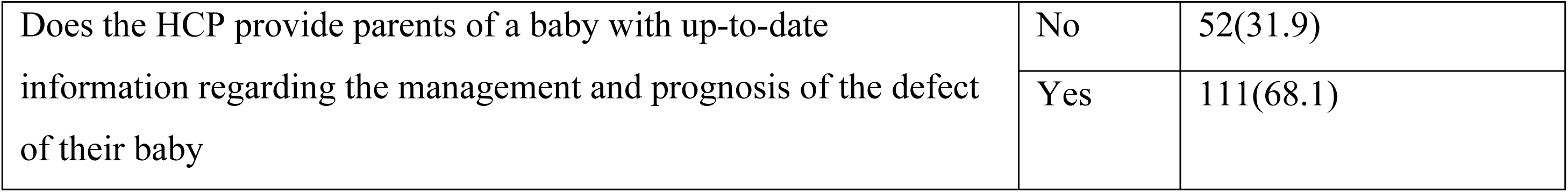
Practice on screening of congenital malformation.

Key findings from practice related items demonstrated suboptimal adherence to comprehensive newborn screening protocols. Only 1.2% of respondents reported the use of a structured surveillance tool for detecting congenital anomalies. While 53.4% routinely performed perineal examinations to assess anal patency, only 48.5% examined the external genitalia, and merely 19.6% inspected the back for signs of neural tube defects. In contrast, pulse oximetry screening was conducted by 76.7% of respondents, indicating comparatively better compliance with recommended practices for the early detection of critical congenital heart defects.

### 5.3 Patient-Related Characteristics

Among the neonates assessed by the participating HCPs, the majority was male, and 46.6% were preterm. Most were born via spontaneous vaginal delivery (62.6%), and only 1.8% were delivered at home. Approximately 22.7% were born after pregnancies complicated by maternal illness or obstetric events.

**Table 5:**
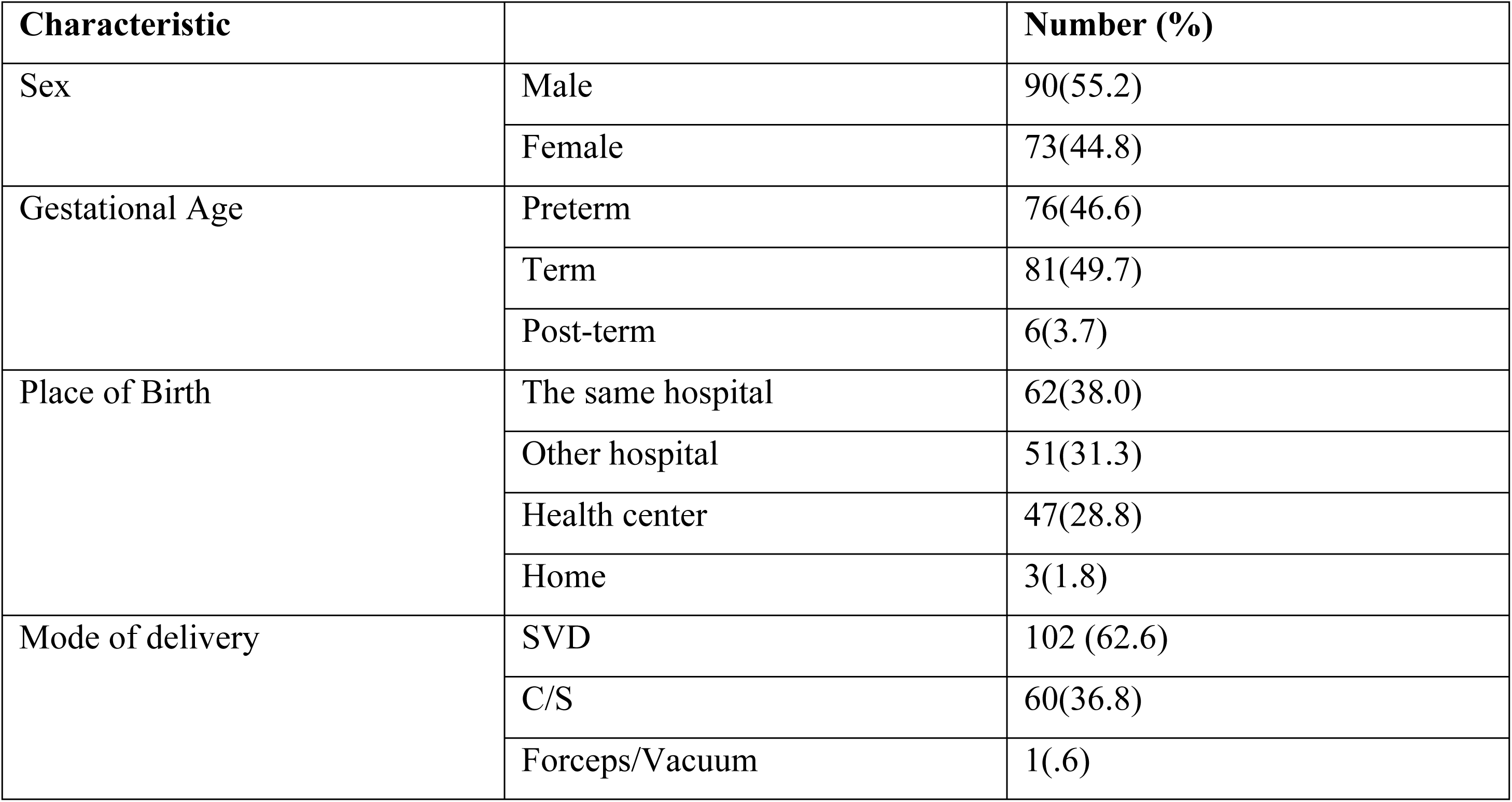
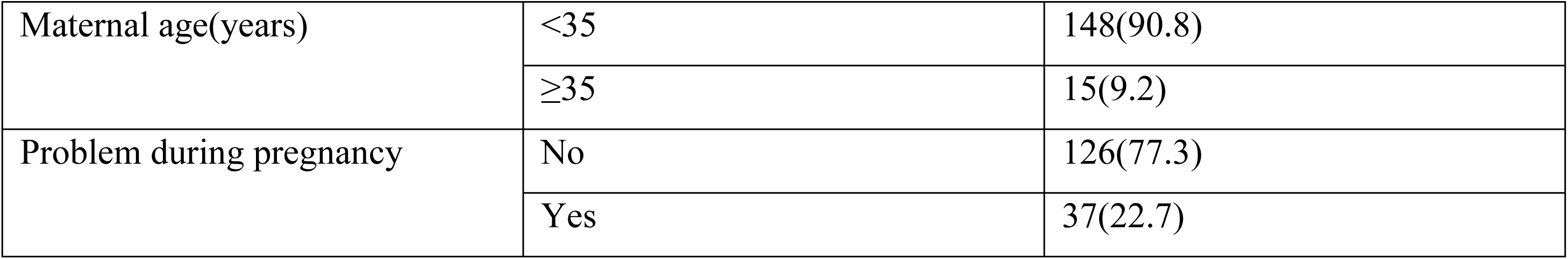
sociodemographic Characteristic of patients.

### 5.4 Facility-Related Factors

Only 6.1% of healthcare providers reported having received formal on-the-job training in newborn screening. Furthermore, 47.9% indicated that there were no available guidelines for congenital anomaly screening in their workplace, while 46% were unaware of the existence of any such guidelines. Only 6.1% confirmed the availability of screening guidelines at their facility, highlighting significant gaps in both training and access to standardized protocols.

### 5.5 Factors Associated with Screening Practice

In the univariate analysis, factors associated with newborn screening practice at a significance level of *p* < 0.25 included knowledge level, attitude, newborn sex, and the presence of maternal complications during pregnancy. In the multivariate logistic regression analysis, adequate knowledge emerged as a strong independent predictor of good screening practice (AOR = 30.84; 95% CI: 0.15–0.95; *p* < 0.001). The presence of maternal complications was also significantly associated with increased likelihood of screening (AOR ≈ 19.10; *p* = 0.01). Additionally, the sex of the newborn influenced screening behavior, with male neonates being nearly 80% more likely to be screened than females (*p* = 0.043). No significant associations were observed with healthcare providers’ age, sex, profession, education level, or years of experience.

**Table 6:**
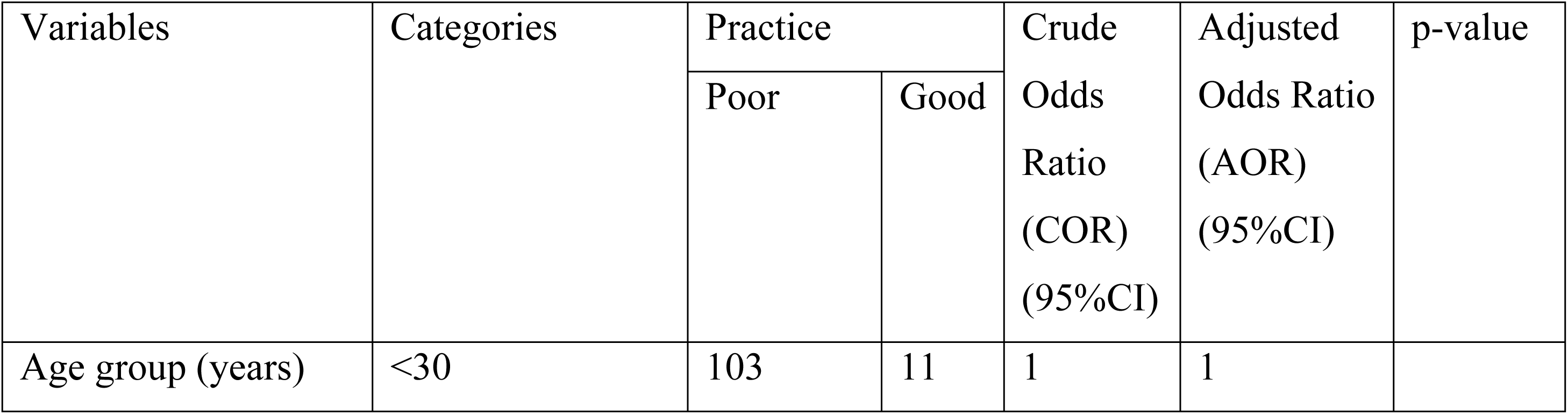
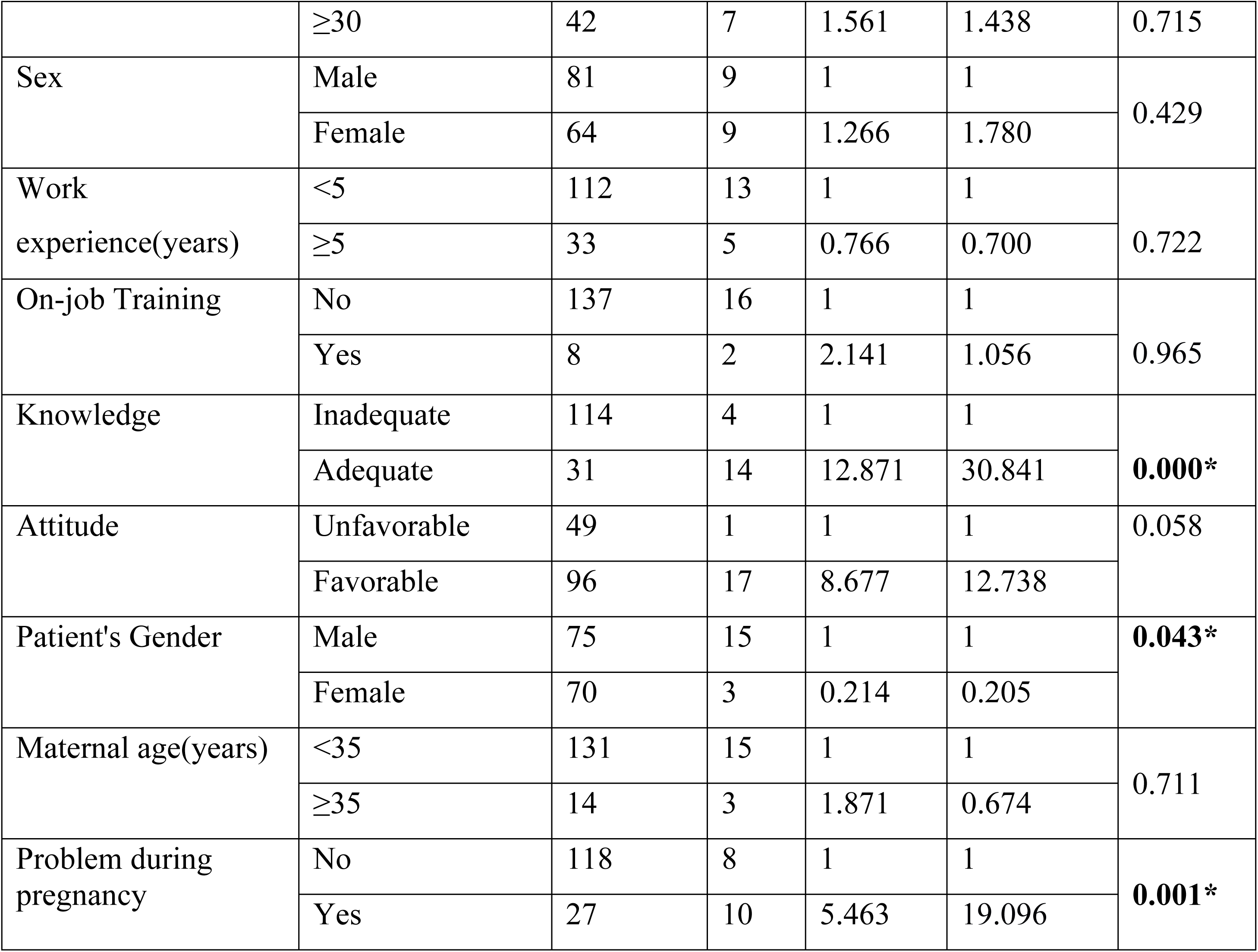
Binomial Logistic Regression analysis results of factors associated with Practice.

| Variables | Categories | Practice |  | Crude Odds Ratio (COR) (95%CI) | Adjusted Odds Ratio (AOR) (95%CI) | p-value |
| --- | --- | --- | --- | --- | --- | --- |
|  |  | Poor | Good |  |  |  |
| Age group (years) | <30 | 103 | 11 | 1 | 1 |  |
|  | ≥30 | 42 | 7 | 1.561 | 1.438 | 0.715 |
| Sex | Male | 81 | 9 | 1 | 1 | 0.429 |
|  | Female | 64 | 9 | 1.266 | 1.780 |  |
| Work experience(years) | <5 | 112 | 13 | 1 | 1 | 0.722 |
|  | ≥5 | 33 | 5 | 0.766 | 0.700 |  |
| On-job Training | No | 137 | 16 | 1 | 1 | 0.965 |
|  | Yes | 8 | 2 | 2.141 | 1.056 |  |
| Knowledge | Inadequate | 114 | 4 | 1 | 1 | **0.000\*** |
|  | Adequate | 31 | 14 | 12.871 | 30.841 |  |
| Attitude | Unfavorable | 49 | 1 | 1 | 1 | 0.058 |
|  | Favorable | 96 | 17 | 8.677 | 12.738 |  |
| Patient's Gender | Male | 75 | 15 | 1 | 1 | **0.043\*** |
|  | Female | 70 | 3 | 0.214 | 0.205 |  |
| Maternal age(years) | <35 | 131 | 15 | 1 | 1 | 0.711 |
|  | ≥35 | 14 | 3 | 1.871 | 0.674 |  |
| Problem during pregnancy | No | 118 | 8 | 1 | 1 | **0.001\*** |
|  | Yes | 27 | 10 | 5.463 | 19.096 |  |

## 6. Discussion

This study assessed the knowledge, attitude, and practice of healthcare professionals regarding the screening of major congenital anomalies among newborns in three tertiary hospitals in Addis Ababa, Ethiopia. The findings reveal critical gaps, particularly in clinical practice, despite moderately high levels of knowledge and favorable attitudes.

Only **11%** of participants demonstrated good screening practice, significantly below international standards. The World Health Organization (WHO) recommends routine and comprehensive newborn screening as a critical step in reducing neonatal morbidity and mortality. The low practice rate observed in this study echoes findings from other low- and middle-income countries (LMICs), where neonatal screening is often inconsistent or neglected (3,14).

In contrast, 27.6% of HCPs had adequate knowledge, and 69.3% demonstrated a favorable attitude. Although better than the practice figures, these numbers still indicate a need for improvement. Adequate knowledge was found to be the strongest independent predictor of good practice, increasing the odds of proper screening by more than thirty times. This underscores the importance of targeted training programs aimed at enhancing provider knowledge.

Additionally, HCPs were more likely to perform screening when the pregnancy was complicated. This likely reflects a risk-based approach to newborn assessment, where complications during pregnancy trigger heightened clinical vigilance. Similarly, male newborns were more frequently screened than females, potentially due to greater attention given to visible genital anomalies such as undescended testes or hypospadias although we ca not conclude from this study.

Only 6.1% of participants reported the availability of institutional guidelines for congenital anomaly screening, and fewer than 10% had received relevant on-the-job training. These institutional deficiencies mirror findings from similar studies in LMICs, where the absence of structured protocols and resource constraints impedes effective neonatal screening (5,6).

Specific practice gaps were identified in key areas of physical examination. While over 75% routinely measured oxygen saturation a promising finding aligned with WHO recommendations for critical congenital heart disease screening, only 19.6% examined the spine for neural tube defects in contrary to a study from Egypt assessing the nurses’ competency level regarding the care of infants with congenital anomalies of central nervous system which showed that 46.55% had competent practice (18).

There is knowledge and practice gap regarding the diagnosis esophageal atresia with or without tracheoesophageal fistula (EA/ TEF) as only 65.6% correctly answered the knowledge question and only 58.2% of HCPs properly assess the newborns for possible chocking episode after initiation of feeding. This finding is consistent with a study done on EA mortality in LMIC is still estimated to vary from 30 to 80% and delay in diagnosis and/or referral of individuals with EA are some of the challenges mentioned in LMIC documented literature. Since aspiration pneumonia has been linked to babies who were referred from outside hospitals, contrast studies, and/or trial of oral feeding, raising pediatricians’ knowledge of EA, cautioning them against performing contrast studies, and developing early referral guidelines (13).

Of the total participants, just 53.4% assessed anal patency. These results are consistent with reports from Pakistan, India and Malawi showing delays in the diagnosis of anorectal malformations, often extending beyond the neonatal period (7,8,10).

In high-income countries, newborn screening is a well-integrated and standardized component of routine neonatal care, contributing significantly to early detection and timely intervention for congenital anomalies. This success is not solely due to the availability of advanced technology, but also to the effective task-sharing among trained healthcare professionals. For instance, studies from the United Kingdom have demonstrated that midwives, when adequately trained, can perform comprehensive newborn assessments with high levels of accuracy and patient satisfaction (15). This highlights an important lesson for low-resource settings like Ethiopia: effective newborn screening does not have to be limited to physicians. Empowering and equipping midwives, nurses, and other frontline health workers through structured training can enhance early detection efforts and bridge current gaps in neonatal care.

Our findings align with Several studies which have identified that there is a knowledge and practice gap among healthcare professionals regarding newborn screening for various abnormalities. Around 50% of family physicians in Kingdom of Saudi Arabia did not screen any child for hearing loss during their last five years of experience (17). In Egypt, 54.45% nurses in a Neurosurgery Department had incompetent total score of practice regarding Congenital anomalies of CNS (18).

This suggests a broader pattern across LMICs where systemic barriers such as training deficiencies and absence of guidelines limit the implementation of best practices.

## 7. Strengths and Limitations

This study has several notable strengths. To our knowledge, it is the first in Ethiopia to assess the knowledge, attitudes, and practices (KAP) of healthcare professionals regarding the screening of major congenital malformations. The multicenter design, involving three tertiary-level hospitals with neonatal intensive care and pediatric surgical services, enhances the representativeness of the findings within the urban referral context. Additionally, the use of direct observation alongside self-reported data strengthened the validity of the practice assessment and helped reduce self-reporting bias. Data collection tools were adapted from internationally recognized WHO and CDC guidelines, ensuring content relevance and alignment with global standards.

However, the study also has limitations. There was no previously validated tool specifically designed to assess KAP related to congenital malformation screening; although instruments were adapted from established guidelines, they have not been externally validated. The study was confined to tertiary public hospitals in Addis Ababa, which may limit generalizability to rural areas or private healthcare settings. While efforts were made to minimize observer bias through the use of anonymous observers and unannounced observation periods, its presence cannot be entirely excluded. Moreover, the study did not evaluate neonatal outcomes, which could have provided additional context for assessing the effectiveness of screening practices.

## 8. Conclusion and Recommendations

### Conclusion

This study found that while the majority of healthcare professionals had moderate to adequate knowledge and a favorable attitude toward screening for congenital anomalies, screening practices were alarmingly poor. The lack of institutional guidelines, inadequate training, and reliance on non-standardized practices contribute to poor practice regarding screening of congenital malformations which in turn causes delayed diagnosis and potentially preventable neonatal morbidity and mortality.

Knowledge level was the most significant factor associated with good screening practice. Additionally, HCPs were more likely to screen infants born after complicated pregnancies or male newborns, suggesting inconsistent application of universal screening principles.

### Recommendations

Based on the findings of this study, we recommend that structured training programs should be implemented for healthcare professionals, focusing on the detection of major congenital anomalies and timely referral. The development and dissemination of standardized national protocols and checklists for neonatal screening across all levels of care are essential to ensure uniformity and adherence. Routine use of structured newborn examination tools including pulse oximetry, perineal inspection, and spinal and genital assessments should be encouraged as standard practice. Strengthening institutional capacity by integrating screening protocols into neonatal and delivery care services, along with supportive supervision and mentorship are also critical. Lastly, further research should be conducted to include rural and community-level settings, as well as studies assessing screening outcomes and the effectiveness of referral pathways.

## List of abbreviations

ARM: anorectal malformation
A.A: Addis Ababa
AAU: Addis Ababa University
AAHB: Addis Ababa health Bureau
CA: Congenital Anomalies
CDC: United States Centers for Disease Control and Prevention
CNS: central nervous system
EA/TEF: esophageal atresia/ tracheoesophageal fistula
HCP: health care professional
LMIC: low and middle income countries
NICU: neonatal intensive care unit
SPHMMC: Saint Paul hospital millennium medical college

## Declarations

### Ethics Approval and Consent to Participate

Ethical approval was obtained from the Institutional Review Board (IRB) of Addis Ababa University, College of Health Sciences. Permission was also secured from the participating hospitals. Written informed consent was obtained from all study participants. Confidentiality and anonymity were maintained throughout the research process.

### Consent for Publication

Not applicable.

### Availability of Data and Materials

The datasets used and analyzed during the current study are available from the corresponding author upon reasonable request.

## Authors’ Contributions

Dr. E.T: study conceptualization and design, data collection and analysis, and drafting and revising the manuscript, coordinated the revision process

Dr. F.T: supervised the research process and provided critical review of the manuscript.

Dr. A. A: Data analysis and interpretation, manuscript writing, review and editing

Dr. N.A: Data collection, Data interpretation, manuscript review and editing

Dr. Y.T: Data collection, Data interpretation, manuscript review and editing

Dr. N.M: Data collection, data validation, manuscript review editing

Dr. T.M: Data collection, manuscript review and editing

Dr. T.L: supervised the research, Data analysis, manuscript review and editing

All authors read and approved the final version of the manuscript.

## Data Availability

All data will be available from Addis Ababa repository https://etd.aau.edu.et/handle/123456789/6718

## Acknowledgments

The authors thank Addis Ababa University and the participating hospitals for their support. Special appreciation is extended to Dr. Abraham, Dr. Netsanet, Dr. Adu, and Dr. Tigist for their assistance in data collection and ongoing support.

